# Red Blood Cell Stiffness Driving Patient Symptoms: A Study of Red Blood Cell Population Rigidity in Sickle Cell Patient Genotype SC Relation to Overlooked Clinical Symptoms

**DOI:** 10.1101/2021.06.11.21257671

**Authors:** Mark Shamoun, Mario Gutierrez, Omolola Eniola-Adefeso

**Affiliations:** Department of Pediatric Hematology/Oncology, University of Michigan, Ann Arbor MI, 48109; Department of Chemical Engineering, University of Michigan, Ann Arbor, MI 48109; Department of Biomedical Engineering, University of Michigan, Ann Arbor, MI 48109; Macromolecular Science and Engineering Program, University of Michigan, Ann Arbor, MI 48109

**Keywords:** Ektacytometry, Deformability Curve, Rigid Red Blood Cells, Sickle Cell Disease, Hgb SS, Hgb SC

## Abstract

Sickle cell disease (SCD) is a systemic hematological disease. Various genotypes of the disease exist; however, the two most common include hemoglobin SS (Hgb SS) and hemoglobin SC (Hgb SC) disease. Hgb SC is typically considered a less severe genotype; however, some patients with SC disease still have significant complications. Ektacytometry is utilized to measure red blood cell deformability in sickle cell patients and may help identify patients at risk for severe disease. We described a patient with genotype hemoglobin SC with a more severe phenotype, who we show to have very rigid red blood cells via ektacytometry.

## Background

Sickle cell disease (SCD), a hereditary multisystem illness, is one of the most prevalent blood cell diseases, affecting over 100,000 Americans and millions worldwide.(1,2) Previous studies have shown many biochemical mechanisms of SCD pathology, including activation of the vascular endothelium, leukocytosis, oxidative stress from tissue reperfusion, and intravascular and extravascular hemolysis.[1] The symptoms of SCD is dependent on the genotype of SCD – two of the most common being, hemoglobin (Hgb) SS and Hgb SC. The Hgb SS is the most prevalent and severe genotype. Simultaenously, the Hgb SC is clinically accepted as milder due to fewer hemolysis markers and fewer associations with severe complications, including stroke, early mortality, and vaso-occlusive pain episodes.(3) However, for both of these subtypes, the red blood cells (RBCs) are often afflicted with increased membrane rigidity due to the high concentration of mutated or abnormal Hgb S inside the RBC. Furthermore, the mutated hemoglobin’s polymerization under hypoxic conditions can alter the RBC shape to an irreversibly-sickled form.

Due to natural aging, the RBC hemoglobin density or mean corpuscular hemoglobin concentration (MCHC) increases moderately; however, this is severely accelerated in SCD.(4–6) During deoxygenation, sickle cells experience a loss of potassium that causes an ion imbalance and the loss of water from the cell, resulting in a higher MCHC.(4,5) Past studies have reported that this loss of potassium from Hgb SC RBCs is higher compared to SS due to the presence of Hgb C.(4) This unique pathophysiology of SC cells results in cells that, although contain approximately half Hgb S compared to SS cells, have a high MCHC. Interestingly, this implies that, depending on their hydration conditions, the SC RBCs can lose membrane deformability as much as, or more, SS cells. (7) However, while they have similar sickling rates to SS cells, the Hgb SC have a reduced capacity to irreversibly sickle – i.e., they are less likely to change shape permanently.(5)

Unsurprisingly, the irreversibly-sickled RBCs have been a primary focus of research in SCD, leading to the understanding that these cells cause significant physical damage when traveling through the body by occluding microvasculature, depriving tissues of nutrients and oxygen, and damaging vital organs such as the spleen, liver, and lungs.(8,9) The combination of vessel occlusion and tissue deprivation of oxygen results in a vaso-occlusive pain crisis experienced by SCD patients. Shape alteration in RBCs is dependent on a variety of factors, including oxygen conditions and hemoglobin contents of individual RBCs. Under stress, Hgb S will spontaneously polymerize and result in RBC shape alteration.(10) Although the staple crescent-like shape is the most frequent shape deformation, it is not always guaranteed.(10–12) However, alterations to the membrane flexibility are certain.(12) Importantly, rigidified, non-deformed RBCs have a longer lifespan than RBCs that have undergone shape deformation to the sickled confirmation.(13,14) Thus, SCD patients live more with the consequence of rigid, normal-shaped, i.e., discocyte, RBCs than the sickle-shaped, which may prove to be more critical in maintaining their general well-being. Herein, we describe a patient with Hgb SC disease with a more severe clinical course, found to have very rigid red blood cells.

## Case Presentation

Our patient of interest (Patient 1) has a history of Hgb SC, first diagnosed at an early age. The patient has one unaffected sibling and another sibling who also has Hgb SC (Patient 2). Over the last few years, Patient 1 has struggled significantly with vaso-occlusive crises (VOC) that seemed exaggerated for their genotype. Over one year, the patient was admitted 12 times for VOC. In the following year, the patient was admitted for 85 days with an average length of stay of 10.6 days. In contrast, patients with Hgb SS studied (n=7) had an average of 1.3 admissions per year and an average length of 5.4 days per stay. For further clinical information regarding Patient 1, contact the corresponding author.

We measured the deformability of blood from several SCD patients in the Erythrocyte Diagnostic Lab (EDL) at Cincinnati Children’s Hospital using an ektacytometer (Lorrca), which quantifies the elongation of RBCs as a function of shear stress at a constant physiologic osmolality. Blood samples were obtained via venipuncture according to a protocol approved by the University of Michigan Internal Review Board (IRB) and following the declaration of Helsinki. Informed consent was obtained for each patient before blood draws, and samples were collected into a syringe containing EDTA as the anticoagulant. TABLE 1 lists all the patients evaluated, including the SCD genotype, general demographics, RBC stiffness measurement, hospitalization, and pain frequency. Blood, shipped to EDL overnight at 4°C, was diluted in 0.14 M polyvinylpyrrolidone to achieve physiological osmolality (∼290 mPa). Healthy, non-SCD blood was used as a control. Except for Patient 1, ektacytometry deformability and Osmoscan analysis were performed once for each patient. The ektacytometry analysis was performed three different times, each ∼16 weeks apart, for Patient 1. All patient samples were collected during routine clinical visits, i.e., collected in a stable state of health.

**TABLE 1:**
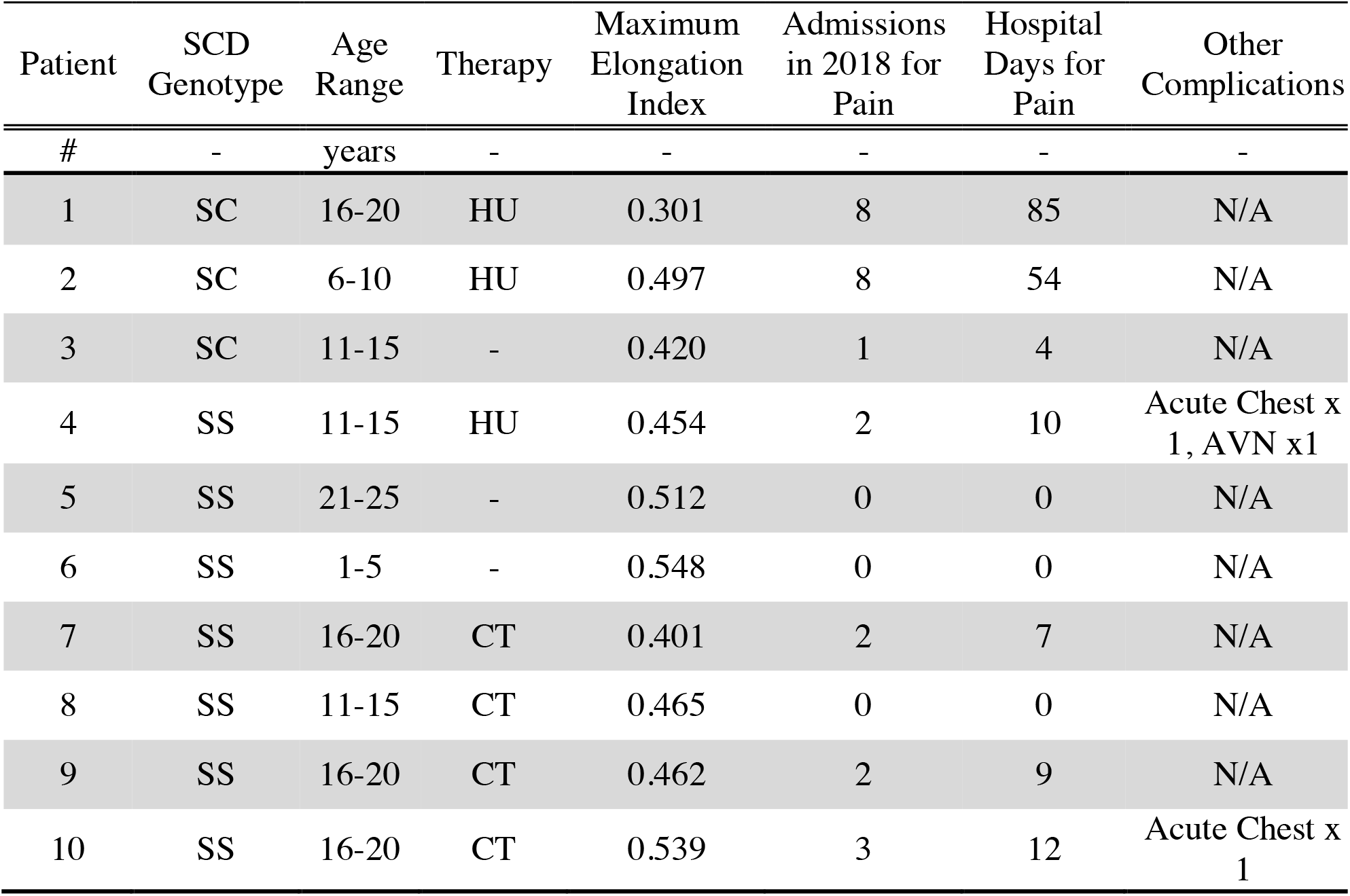
Sickle Cell Disease Patient and Hospitalization information. Table with patient information including SCD genotype, age range in years, Elongation Index recorded via ektacytometry of the patient blood sample, and current patient therapy chronic transfusions (CT) or Hydroxyurea (HU). Missing therapies indicate the patient is not under any type of treatment at the time of the measurement. The table also includes Information regarding the patient hospitalization frequencies and medical complications.

| Patient | SCD Genotype | Age Range | Therapy | Maximum Elongation Index | Admissions in 2018 for Pain | Hospital Days for Pain | Other Complications |
| --- | --- | --- | --- | --- | --- | --- | --- |
| # | - | years | - | - | - | - | - |
| 1 | SC | 16-20 | HU | 0.301 | 8 | 85 | N/A |
| 2 | SC | 6-10 | HU | 0.497 | 8 | 54 | N/A |
| 3 | SC | 11-15 | - | 0.420 | 1 | 4 | N/A |
| 4 | SS | 11-15 | HU | 0.454 | 2 | 10 | Acute Chest x 1, AVN x1 |
| 5 | SS | 21-25 | - | 0.512 | 0 | 0 | N/A |
| 6 | SS | 1-5 | - | 0.548 | 0 | 0 | N/A |
| 7 | SS | 16-20 | CT | 0.401 | 2 | 7 | N/A |
| 8 | SS | 11-15 | CT | 0.465 | 0 | 0 | N/A |
| 9 | SS | 16-20 | CT | 0.462 | 2 | 9 | N/A |
| 10 | SS | 16-20 | CT | 0.539 | 3 | 12 | Acute Chest x 1 |

Figure 1 shows the deformability curve obtained for all patient blood evaluated with Figure 1A showing RBC elongation analysis at constant osmolality and variable shear stress. Figure 1B shows elongation analysis with constant shear and variable osmolality, i.e., osmoscan. From these curves, a maximum Elongation Index (EI_MAX_) can be defined as the maximum deformability of RBCs at high shear stress, which is accepted as a measure of the rigidity in blood. A lower EI_MAX_ value relative to healthy blood is indicative of the presence of stiff RBCs. As with previous reports, the estimated EI_MAX_ for healthy blood is ∼0.63.(15) The EI_MAX_ obtained for Patient 1 was 0.30, while the blood from other patients ranged from 0.40 to 0.55.

**Figure 1:**
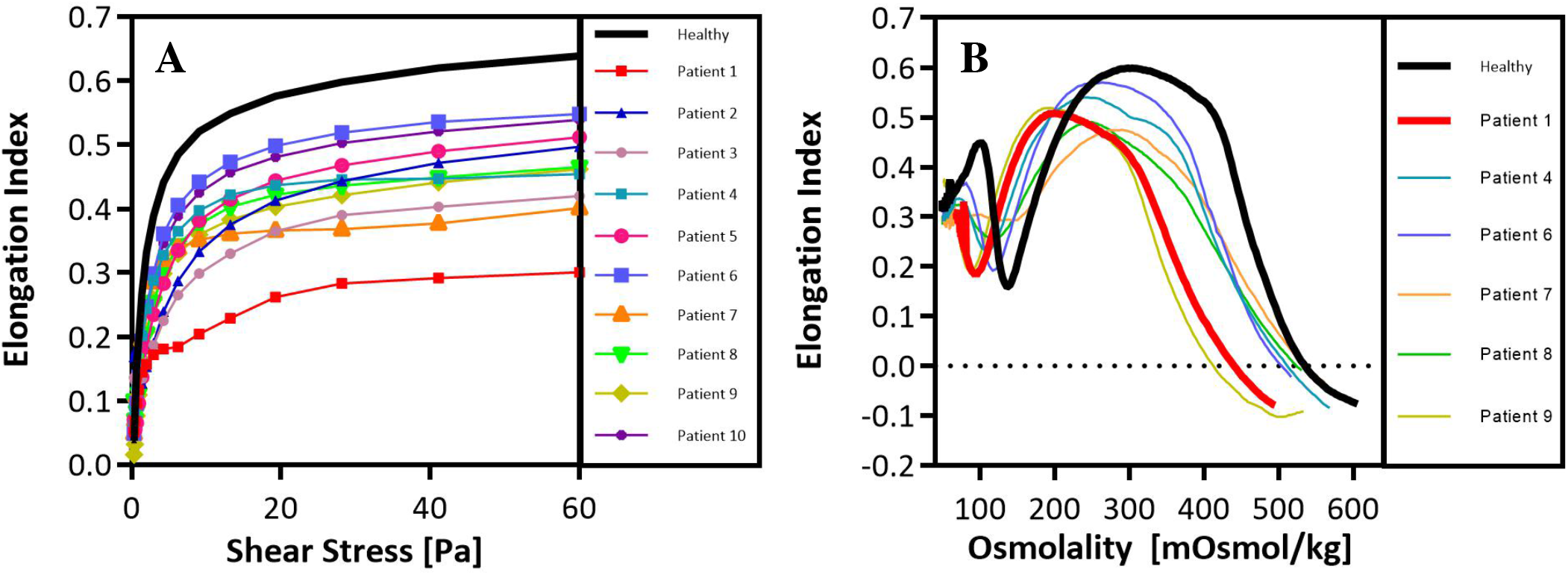
Sickle Cell Patient Red Blood Cell Deformability analysis via Ektacytometry. A, Standard ektacytometry deformability analysis at constant osmolality (∼290 mOsmol/kg) and variable shear stress. B, Osmoscan ektacytometry analysis at constant shear stress (∼30 Pa) and variable osmolality.

We find a moderate negative Pearson correlation between the EI_MAX_ values measured and the number of hospital days for pain. There is a very significant difference between the EI_MAX_ values measured from Patient 1 (*μ*_pat1_=0.332, n=3) and the EI_MAX_ values of the other nine patients (*μ*_ops_=0.478, n=9), yielding a p-value of 0.003 as determined by a Student’s t-test 1-tail analysis. Indeed, the mean elongation of Patient 1 blood is classified as an outlier when grouped with the other nine patients’ elongations. By excluding Patient 2 from this analysis, we find a very significant difference (p=0.009) for the number of hospital days for pain per admission between Patient 1 and Patients 3-10. In fact, for Patients 3-10, there is an average of 4.2 hospital days with pain for every admission, compared to the 10.6 hospital days with pain per admission for Patient 1.

## Discussion & Conclusions

Previous studies have used ektacytometry to measure rigidity as a function of changing osmolality at constant shear stress.(16) Figure 1A shows a comparison of our deformability data at constant osmolality with varying shear stress and vise-versa in Figure 1B. We opted to interpret our EI_MAX_ with a deformability analysis based on data collected at constant osmolality and varied shear stress rather than with osmoscan since the former mimics the physiologic environment where RBCs experience changing shear stress as they flow through the various blood vessels at a fixed, physiologic osmolality. As shown, patient 1, which is our patient described above, showed the lowest EI_MAX_ (∼0.301) indicative of an ultra-rigid RBC population, which is impressive given his SC genotype but fits with the high VOC experienced by this patient. Similar to observations seen by Ballas *et al*., we observe from Figure 1B that the RBCs in the blood sample from Patient 1 are highly dehydrated, as expected to be seen in an SC sample.(7) This dehydration is likely one of the factors driving the extreme rigidity of the RBCs in the patient.

Our patient and his sibling (Patient 2) were on hydroxyurea treatment in 2017 through 2019, known to increase RBC deformability and a resulting higher EI_MAX_ value.(16) However, despite hydroxyurea treatment, our patient still had a reduced EI compared to other patients. In 2019, our patient was started on chronic transfusions with a partial exchange, given his severe phenotype. After initiating chronic transfusions, his EI_MAX_ increased (∼0.37), and the frequency of pain crisis subsequently decreased, as shown in Figure 2, Measurement 3. Patients 5 and 6 show similar curves to our healthy control, and neither required hospitalization for sickle cell disease complications. Patient 10 did have multiple hospitalizations; however, this patient did have difficulty with transportation, and although they were on chronic transfusions, appointments were missed. Patient 10’s rigidity was obtained while the patient was on every 3 to 4-week transfusions, and the missed appointments likely contributed to the hospitalizations.

**Figure 2.**
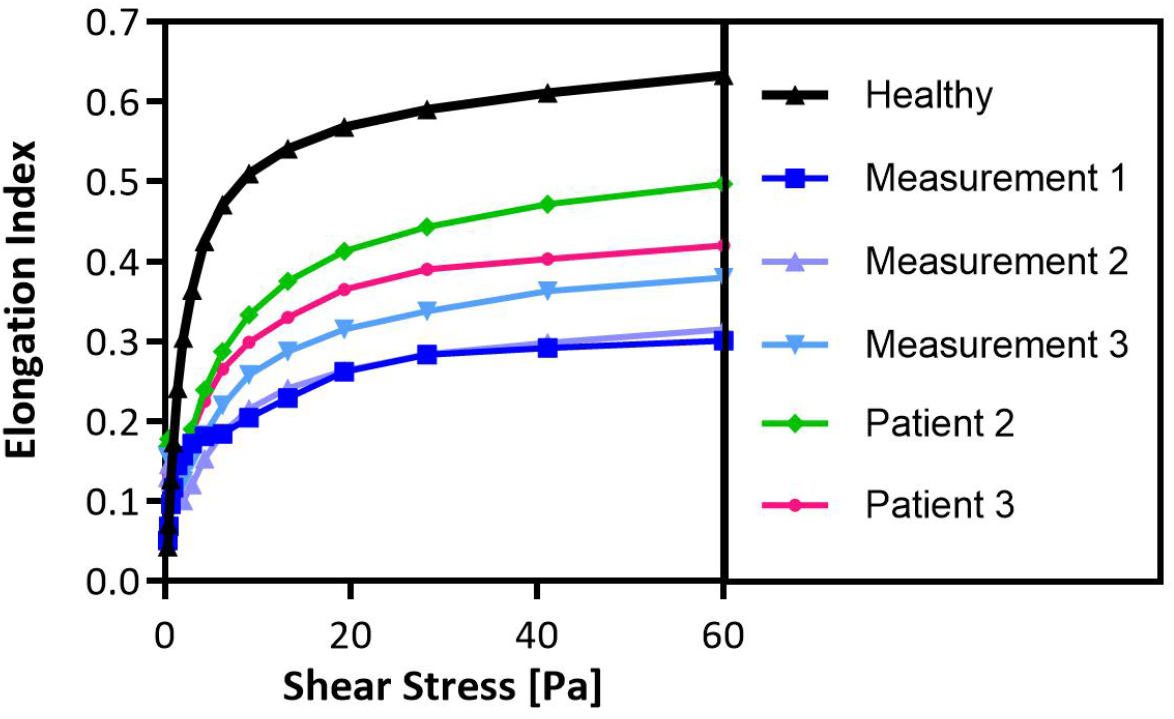
Deformability analysis via Ektacytometry of Focus Patient in Comparison to other SC Patients. Standard ektacytometry deformability analysis at constant osmolality (∼290 mOsmol/kg) and variable shear stress. Ektacytometry analysis of focus patient taken at three different time periods, compared to deformability analysis of other SC patients.

The patient’s younger sibling was also analyzed (Patient 2); however, their EI_MAX_ was higher and closer to other SCD patients’ values. Clinically, his sibling still required multiple admissions, but had 54 hospital days in 2018 and required significantly less intervention per stay. During many of the admissions, Patient 2 did not require a PCA and instead could be managed on intermittent intravenous morphine or oral pain medications. Previous studies focusing on the psychological impact of SCD on children and their siblings suggest little attention is given to family members or siblings who are not chronically ill or are healthy.(17–19) These siblings often are subject to anxiety, feelings of rejection, and even jealousy.(17–19) Given the context of this complicated social situation, it is likely that the number of hospital days and admissions of Patient 2 are skewed and do not represent an independent patient clinical outcome. For these reasons, we opted to remove Patient 2 from a statistical analysis comparing the hospital days with pain per admission between all the patients (Patients 3-10) and Patient 1.

Additionally, we compare Patient 1 to Patient 3, Figure 2. Patient 3 is also an SC genotype and expresses a phenotype much more typically seen in SC patients. The EI_MAX_ observed for Patient 3 is 0.42, closer to values observed in stable patients. In comparison, Patient 3 was admitted only once in 2018 for four days with pain. Certainly, Patient 3’s clinical stability is something regularly observed in SC patients.

In conclusion, our patient is observed to have abnormally highly rigid RBCs despite treatment and presents an abnormally high pain occurrence for an SCD patient with what is regarded as a clinically less severe genotype, SC. Previous studies show SC patients tend to receive a late diagnosis, often not until a serious or fatal health complication.(20) Utilizing RBC deformability analysis may allow for identifying patients with high risk for complications in the absence of a significant level of hemolysis or irreversible sickling. Although it is difficult to determine the entire and exact contribution RBC rigidity has on VOC’s development in our patient, it is likely a contributing factor to the variation in patient disease phenotypes in SCD. It warrants further exploration in how one could use RBC deformability analysis for predictive outcomes in patients, especially for those not on chronic transfusions or hydroxyurea therapies.

## Materials & Methods

### Human Study Approvals

This study was reviewed and received approval from the University of Michigan Internal Review Board (IRB-MED and IRB-HSBS). All procedures were conducted following the tenets of the Declaration of Helsinki. Study subjects and parents were informed of study protocols and agreed to informed consent before blood collection.

### Preparation of SCD Patient Blood

Fresh blood was obtained on the day of a patient’s routine clinical visit via venipuncture. Blood was drawn into standard Vacutainer Lavender K2-EDTA tubes (BD) and stored at 4 ºC. Blood samples were shipped in 4 ºC cold packs overnight to Erythrocyte Diagnostics Laboratory of the Cancer and Blood Diseases Institute at the Cincinnati Children’s Hospital for analysis the following day. All SCD patient samples were measured within 24 hours of blood draw. Complete blood counts were run on a Sysmex 9100 XN automated machine. The sample was then sent to the hematology lab at Michigan Medicine and run according to university protocol and manufacturer’s instructions. Results were verified and reported through the electronic medical records at the University of Michigan.

### Ektacytometry Deformability Measurements

All samples were measured independently using a LoRRca Maxsis Ektacytometer (Mechatronics Instruments BV, Zwaag, The Netherlands) at the Erythrocyte Diagnostics Laboratory of the Cancer and Blood Diseases Institute at the Cincinnati Children’s Hospital. SCD whole blood samples were diluted ∼200x in polyvinylpyrrolidone (MW 360,000) solution. The solution is then transferred into LoRRca automized measuring vessel. Measurements are taken through a range of shear stresses (Pa) up to a maximum of 60 Pa. Cell deformation is expressed as an elongation index calculated by equation (1), where A represents the major axis and B the minor axis in deformation. Elongation Index (EI) values are plotted versus shear stress (Pa) to obtain the deformability curve. The osmoscans sample was prepared in an identical method.

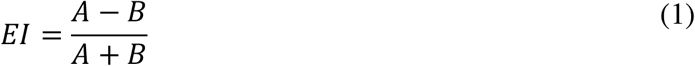

## Data Availability

All data generated or analyzed during this study are included in this published article and supplementary material. The raw datasets in this current study are available from the corresponding author on reasonable request. The identity of study participants will not be shared.

## Abbreviations

SCD: Sickle Cell Disease
HgB: Hemoglobin
RBC: Red Blood Cells
MCHC: Mean Corpuscular Hemoglobin Concentration
VOC: Vaso-Occlusive Crises
PCA: Patient Controlled Analgesia
EDL: Erythrocyte Diagnostic Laboratory
IRB: Internal Review Board
EDTA: Ethylenediaminetetraacetic Acid
EI: Elongation Index

## Declarations

### Ethics Approval and Consent to Participate

Blood draw protocols have been approved by the University of Michigan Internal Review Board (IRB-MED). SCD donors and legal guardians are informed of the study and give written consent before blood collection. Fresh blood is obtained on the day of a patient’s routine clinical visit via venipuncture. Blood is drawn into standard Lavender Vacutainer K2-EDTA for ektacytometry and hemoglobin electrophoresis analysis. Blood sample for ektacytometry analysis, lavender tube, was stored and shipped in 4 ºC cold packs overnight to Erythrocyte Diagnostics Laboratory of the Cancer and Blood Diseases Institute at the Cincinnati Children’s Hospital for examination the following day. All SCD patient samples are measured within 24 hours of blood draw.

### Consent for Publication

SCD donors and legal guardians are informed of the study and give written consent for publication. Identity of donors and relatives has been removed for publication purposes.

### Availability of Data

All data generated or analyzed during this study are included in this published article and supplementary material. The raw datasets in this current study are available from the corresponding author on reasonable request. Identity of study participants will not be shared.

### Competing Interests

The authors declare no competing interests.

### Funding

This work was in part supported by an NSF Research Grant (CBET1854726) to O.E.A. M.G. was supported by National Science Foundation Graduate Research Fellowship.

### Author Contributions

M.S. and O.E-A conceived and designed the experimental protocols. M.G. prepared all blood samples for measurement and analyzed all the collected data. M.G., M.S., and O. E-A wrote the manuscript. M.S. interacted with all patients as a clinician, including reading consents to donate and blood collection and provided valuable clinical insight. All authors read and approved the final manuscript.

## Acknowledgements

The authors thankfully acknowledge the patients and families for their participation in this research study.

## Notes

### Competing Interest Statement

The authors have declared no competing interest.

### Author Declarations

Blood draw protocols have been approved by the University of Michigan Internal Review Board (IRB-MED). SCD donors and legal guardians are informed of the study and give written consent before blood collection.

